# Epigenetic clocks are not accelerated in COVID-19 patients

**DOI:** 10.1101/2020.11.13.20229781

**Authors:** Julia Franzen, Selina Nüchtern, Vithurithra Tharmapalan, Margherita Vieri, Miloš Nikolić, Yang Han, Paul Balfanz, Nikolaus Marx, Michael Dreher, Tim H. Brümmendorf, Edgar Dahl, Fabian Beier, Wolfgang Wagner

## Abstract

Age is a major risk factor for severe outcome of coronavirus disease 2019 (COVID-19), but it remains unclear if this is rather due to increased chronological age or biological age. During lifetime, specific DNA methylation changes are acquired in our genome that act as “epigenetic clocks” allowing to estimate donor age and to provide a surrogate marker for biological age. In this study, we followed the hypothesis that particularly patients with accelerated epigenetic age are affected by severe outcomes of COVID-19. Using four different age predictors, we did not observe accelerated age in global DNA methylation profiles of blood samples of nine COVID-19 patients. Alternatively, we used targeted bisulfite amplicon sequencing of three age-associated genomic regions to estimate donor-age of blood samples of 95 controls and seventeen COVID-19 patients. The predictions correlated well with chronological age, while COVID-19 patients even tended to be predicted younger than expected. Furthermore, lymphocytes in nineteen COVID-19 patients did not reveal significantly accelerated telomere attrition. Our results demonstrate that these biomarkers of biological age are therefore not suitable to predict a higher risk for severe COVID-19 infection in elderly patients.

## Introduction, Results and Discussion

Coronavirus disease 2019 (COVID-19) with severe or even fatal outcome disproportionately affects elderly people (Mueller et al., 2020). The infection fatality rate (IFR) exponentially increases with age: while the IFR is zero in children it starts to increase at approximately 55 years (0.4 %) and finally leads to an IFR of 15 % in 85 year old adults (Levin et al., 2020). Chronological age is thus one of the major risk factors to develop severe symptoms during an infection with severe acute respiratory syndrome coronavirus 2 (SARS-CoV-2) and this risk is independent of other age-related comorbidities like diabetes, cardiovascular diseases, obesity (Mueller et al., 2020), or the detection of clonal hematopoiesis of indeterminate potential (CHIP) (Hameister et al., 2020). Recently, it has been suggested that particularly elderly people with accelerated biological age are susceptible to severe disease outcomes (Kuo et al., 2020).

The process of biological aging is reflected by molecular hallmarks of aging, which include epigenetic modifications (Lopez-Otin et al., 2013). There are highly reproducible DNA methylation changes during aging that are acquired at specific CG dinucleotides, so called “CpG sites”. The DNA methylation levels of several age-associated CpGs can therefore be combined into “epigenetic clocks” to predict donor age (Bell et al., 2019; Koch and Wagner, 2011). Notably, various diseases have been associated with accelerated epigenetic aging and accelerated epigenetic age predictions for blood samples are indicative for higher all-cause mortality (Marioni et al., 2015). Thus, epigenetic age of blood does not only reflect chronological age but also aspects of biological age (Field et al., 2018). We therefore followed the hypothesis that accelerated epigenetic age increases susceptibility to severe COVID-19 infections (Mueller et al., 2020; Santesmasses et al., 2020).

In this study, we used blood samples of 20 patients with severe SARS-CoV-2 infection either with or without acute respiratory distress syndrome (ARDS; Supplemental Table S1). All samples were collected at the University Hospital of RWTH Aachen, after written informed consent, and were provided by RWTH cBMB, the biobank of the Medical Faculty of RWTH Aachen University. The mean age was 66 years (32 – 85 years) and 13 of the 20 samples suffered from ARDS.

Since DNA methylation profiles of COVID-19 patients have so far hardly been addressed, the initial nine samples were tested with the Illumina EPIC methylation microarray that investigates approximately 850,000 CpG sites across the genome as part of a pilot study. For comparison we utilized publicly available DNA methylation profiles of 185 healthy blood control samples, which were generated before the first outbreak of SARS-CoV-2. A direct comparison of these studies was hampered by different Illumina BeadChip platforms (450k and EPIC) and batch effects, which despite quantile normalization become apparent in principle component analysis (PCA; Supplemental Figure S1). However, despite batch effects, the results of Illumina BeadChip profiles usually provide relatively robust results for epigenetic signatures, which are based on multiple CpGs (Han et al., 2020; Horvath, 2013; Lin et al., 2016). For orientation, we initially estimated the cellular composition of leucocytes based on DNA methylation profiles, as described by Houseman et al. (2012). We observed significantly lower predictions for CD4 T cells in COVID-19 patients as compared to controls (p = 0.0024, Welch’s t-test; Supplemental Figure S2A), which is in line with results of other groups that investigated leukocyte subsets by flow cytometry (Diao et al., 2020). Furthermore, for our 18 COVID-19 samples with available blood counts the percentage of lymphocytes was significantly lower than in healthy control samples (p < 10^−08^, Welch’s t-test; Supplemental Figure S2B).

We used four different epigenetic age predictors for the Illumina BeadChip profiles: 1) a frequently used aging signature that has been trained for multiple tissues by Horvath (2013) with 353 CpGs; 2) a more recent skin and blood clock of Horvath et al. based on 391 relevant CpGs (Horvath et al., 2018); 3) a frequently used aging signature by Hannum et al. (2013), which was trained on blood samples and utilizes 71 CpGs; and 4) our recently described age-predictor for blood that was trained on 65 CpGs (Han et al., 2020). Signatures 1 and 3 comprise CpGs that were not measured by the EPIC BeadChip (19 and 6, respectively), and this might result in a moderate offset of age-predictions. Nevertheless, with all four signatures the epigenetic age-predictions of the nine COVID-19 samples correlated clearly with chronological age (Figure 1A) and there was a mean absolute error of 6 years, 5 years, 7 years, and 6 years, respectively. Notably, in comparison to controls the COVID-19 samples did not reveal accelerated epigenetic age: the difference between epigenetic age predictions and chronological age, referred to as “delta age”, centered around zero years with the different predictors (Figure 1B).

**Figure 1.**
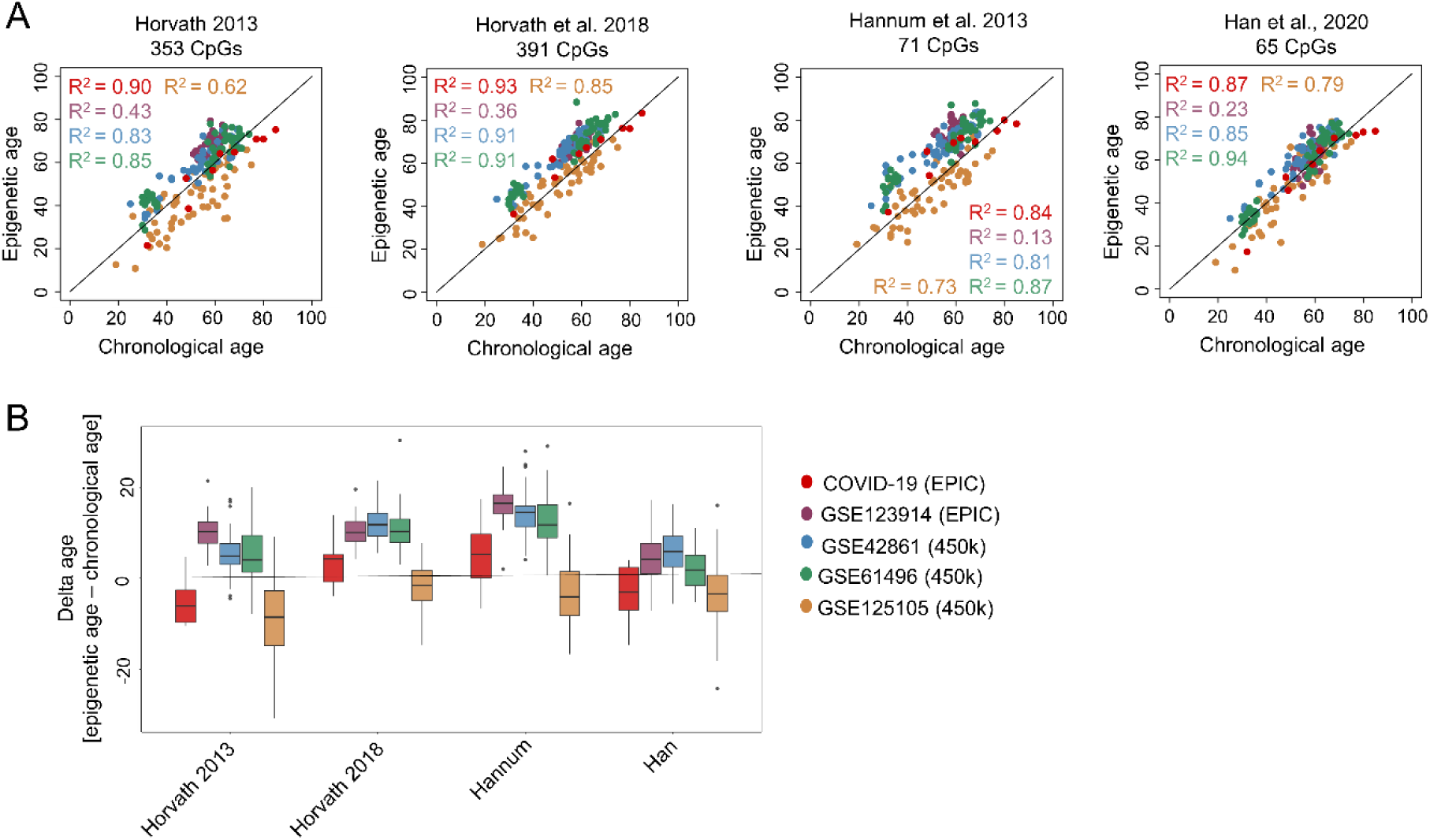
Epigenetic ageing clocks based on Illumina BeadChip data are not accelerated in COVID-19 patients. **A)** Genome wide DNA methylation profiles of nine COVID-19 patients (Illumina EPIC array) and 185 healthy controls of publicly available datasets (EPIC or 450k array) were analyzed with four different predictors of epigenetic age as described by Horvath (2013), Horvath et al. (2018), Hannum et al. (2013) and Han et al. (2020). **B)** Boxplots present the deviation of epigenetic age prediction and chronological age (delta age). Overall, the DNA methylation profiles of the nine COVID-19 samples (indicated in red) did not show consistent age-acceleration across the different aging signatures.

Subsequently, we tested these and additional COVID-19 samples with a targeted bisulfite amplicon sequencing (BA-seq) of age-associated regions. We have recently described BA-seq for nine CpGs that provide robust and reliable age predictions (Han et al., 2020). To further ease applicability of the method we have meanwhile refined the signature to focus on the three regions with highest correlation with chronological age and with a combination of hyper- and hypomethylated CpGs to reduce the PCR bias. The three relevant CpG sites are associated with the genes Coiled-Coil Domain-Containing Protein 102B (*CCDC102B*), Four And A Half LIM Domains Protein 2 (*FHL2*) and Phosphodiesterase 4C (*PDE4C*), as described before (Han et al., 2020). We utilized BA-seq data of 40 healthy blood samples of our previous work to derive the following multivariable model:

Epigenetic age (years) = −0.34 DNAm^CCDC102B^ + 0.83 DNAm^FHL2^ + 1.18 DNAm^PDE4C^ + 3.86

We initially validated this method with 78 blood samples of healthy donors (18 – 83 years) that were collected before the begin of the SARS-CoV-2 pandemic. The epigenetic age predictions revealed a mean absolute error of 4.23 years and a coefficient of determination with chronological age of R^2^ = 0.83. Subsequently, we processed blood samples of 17 COVID-19 patients in parallel with 17 age-matched healthy controls (Figure 2A). There was no evidence for accelerated epigenetic aging in the COVID-19 samples, even if we stratified into samples with or without ARDS (Figure 2B). There was also no evidence for accelerated DNA methylation changes when we analyzed the age-associated CpGs individually (Supplemental Figure S3).

**Figure 2.**
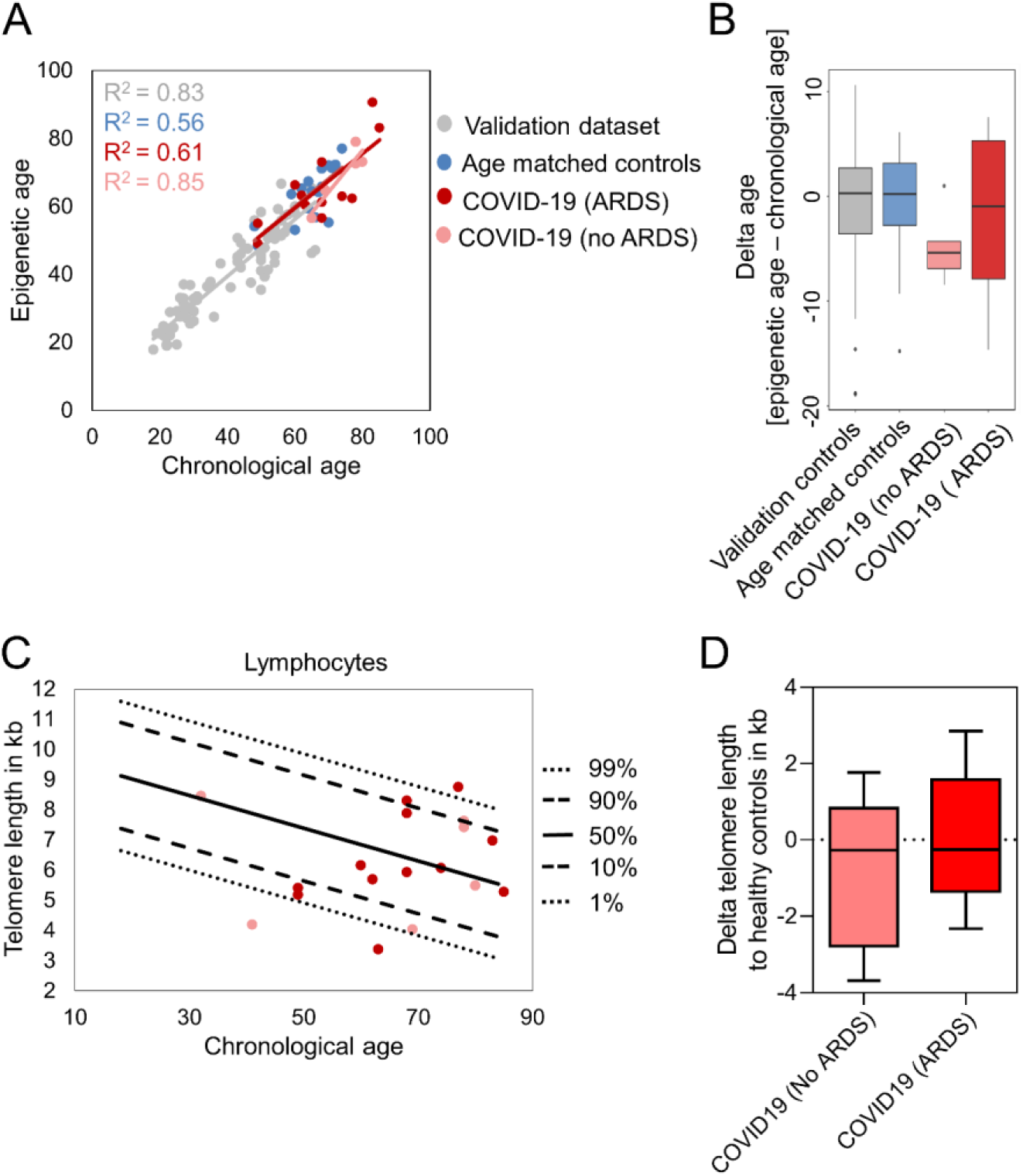
Epigenetic age predictions with amplicon sequencing and telomere length analysis in COVID-19 patients. **A)** Three age-associated CpGs in the genes *CCDC102B, FHL2*, and *PDE4C* were analyzed with targeted bisulfite amplicon sequencing (BA-seq). A multivariable model for epigenetic age-predictions was validated with 78 controls (grey). Subsequently, we analyzed 17 blood samples of COVID-19 patients with (n = 12, red) or without ARDS (n = 5, light red) and 17 age-matched controls (blue). **B)** The deviation of chronological and predicted epigenetic age (delta-age) is presented (no significant differences; Welch’s t test). **C)** Telomere lengths in kilo bases of 19 COVID-19 patients. Lines indicate percentiles of the telomere lengths of 356 healthy controls. **D)** The telomere length difference of no ARDS and ARDS patients to the respective age-adapted telomere length.

Telomere attrition is another hallmark of aging and correlates with age in peripheral blood cells (Brümmendorf and Balabanov, 2006). A recent study indicated that in leukocytes of COVID-19 patients telomere attrition below the 10^th^ percentile is more frequent than in healthy controls (Froidure et al., 2020), however the results of this study might be affected by the lymphopenia observed in COVID-19 patients (Benetos et al., 2020). To further investigate if this alternative biomarker for biological age is accelerated, we analyzed telomere length in lymphocytes of 19 COVID-19 patients with Flow-FISH, as described in detail before (Ferreira et al., 2020; Kirschner et al., 2020). In comparison to the telomere length distribution of 356 healthy controls (Werner et al., 2015) there was overall no evidence for significant telomere attrition in COVID-19 patients (Figure 2C,D).

Taken together, despite of the limitations of a rather small sample size, our results do not provide evidence that severe outcome of COVID-19 is associated with accelerated epigenetic age or significantly shortened telomere length. Thus, we did not observe signs of premature biological aging in our COVID-19 patients. On the other hand, the infection with SARS-CoV-2 did not directly impact on epigenetic age-predictions. It has recently been demonstrated that HIV infection leads to an average aging advancement of 4.9 years, linking molecular aging, epigenetic regulation and disease progression in this retroviral disease (Gross et al., 2016). Furthermore, coronavirus infections have been suggested to mediate DNA methylation at antigen-presentation-associated gene promoters (Menachery et al., 2018). It is still unclear if the different tissues of a human organism reveal the same pace of epigenetic aging and hence it is conceivable that nasopharyngeal and bronchial epithelium, which is preferentially infected by SARS-CoV-2, reveals higher delta-age with epigenetic age-predictions. A recent study suggested that in airway epithelial cells of healthy controls there is age-associated hypomethylation at a CpG site (cg08559914) located near the transcription start site of the receptor angiotensin-converting enzyme 2 (*ACE2*) that permits cell entry of SARS-CoV-2 (Corley and Ndhlovu, 2020). Thus, specific age-associated DNA methylations may be functionally relevant to increase susceptibility to fatal COVID-19. A follow up study with a larger sample size would be required to ultimately rule out a predictive value of either telomere length or age-associated methylation changes in defined subpopulations of patients affected by COVID-19. Either way, our results demonstrate that age-associated DNA methylation changes are not generally accelerated in patients with severe SARS-CoV-2 infections. Analysis of epigenetic age in blood is therefore not suitable to stratify elderly patients that are potentially even more susceptible to severe COVID-19 infections.

## Data Availability

All raw data will be provided upon reasonable request.

## Acknowledgements

This work was particularly supported by the German Research Foundation (to WW: WA 1706/8-1 and WA 1706/12-1 within the CRU344; to NM: TRR 219; Project-ID 322900939 [M03, M05]); the Interdisciplinary Center for Clinical Research within the faculty of Medicine at the RWTH Aachen University (WW: O3-3), Deutsche Krebshilfe (WW: TRACK-AML); and the Federal Ministry of Education and Research (WW: VIP + Epi-Blood-Count). The COVA (COVID-19-Aachen) Study was initiated and financed by the University Hospital Aachen.

## Author contributions

JF contributed to experimental design, data analysis and writing of the manuscript; SN and VT performed MiSeq experiments and MN supported the analysis; MV,FB and THB measured telomere length; YH established the 3CpG ageing model; ED, PB, NM and MD provided blood, DNA samples and data of COVID-19 patients; WW contributed to experimental design, data analysis and writing of the manuscript. All authors revised and approved the final manuscript.

## Supplemental Material

### Supplemental_Material.pdf

Combined PDF with Supplemental Figures S1, S2 and S3, Supplemental Table S1, and Supplemental Methods.

