## Supplemental Figures S1, S2 and S3, Supplemental Table S1, and Supplemental Methods for "Epigenetic clocks are not accelerated in COVID-19 patients"

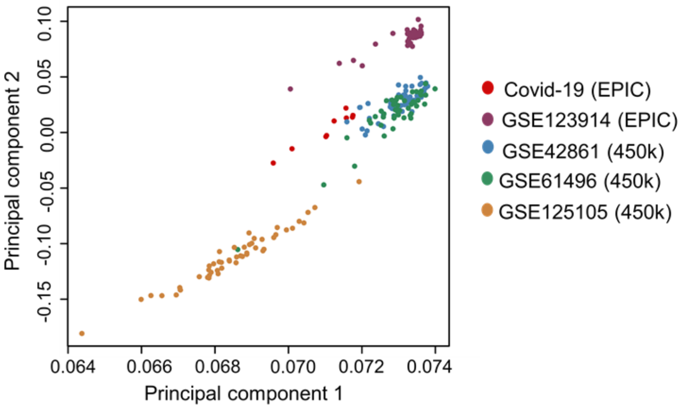


**Supplemental Figure S1. Principal component analysis of DNA methylation microarray datasets.**

Nine whole blood samples of COVID-19 patients have been investigated by the Illumina EPIC microarray and compared to 185 controls of four studies (accession numbers for the Gene Expression Omnibus are provided; https://www.ncbi.nlm.nih.gov/geo/; for better comparison we randomly selected up to 50 DNAm profiles per study). We only considered CpG sites that are represented on both platforms (Illumina EPIC or 450k BeadChips) and excluded CpGs on Y and X chromosome. Despite quantile normalization the samples still cluster according to the different studies.

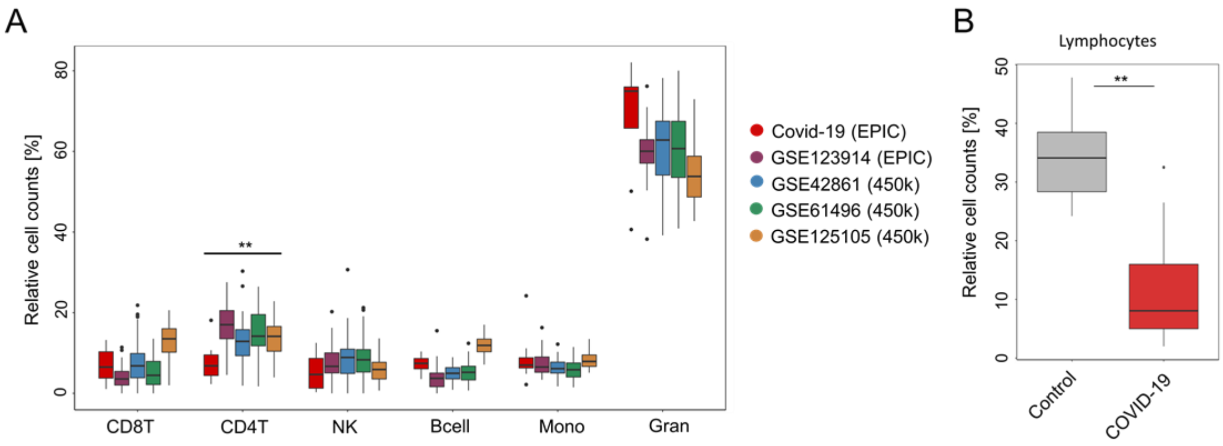


**Supplemental Figure S2. Leukocyte counts.**

**A)** The cellular composition of leukocyte subsets was estimated based on DNA methylation profiles in nine COVID-19 patients and 185 healthy controls from publicly available datasets using the predictor of Houseman et al. (2012). Welch's t test demonstrated a significantly lower fraction of CD4 T cells in COVID-19 patients (** p-value = 0. 0024). **B)** Automated cell counting shows a significant decrease of lymphocytes in 18 COVID-19 samples in comparison to 16 healthy controls (** $p < 10^{-08}$, Welch's t test).

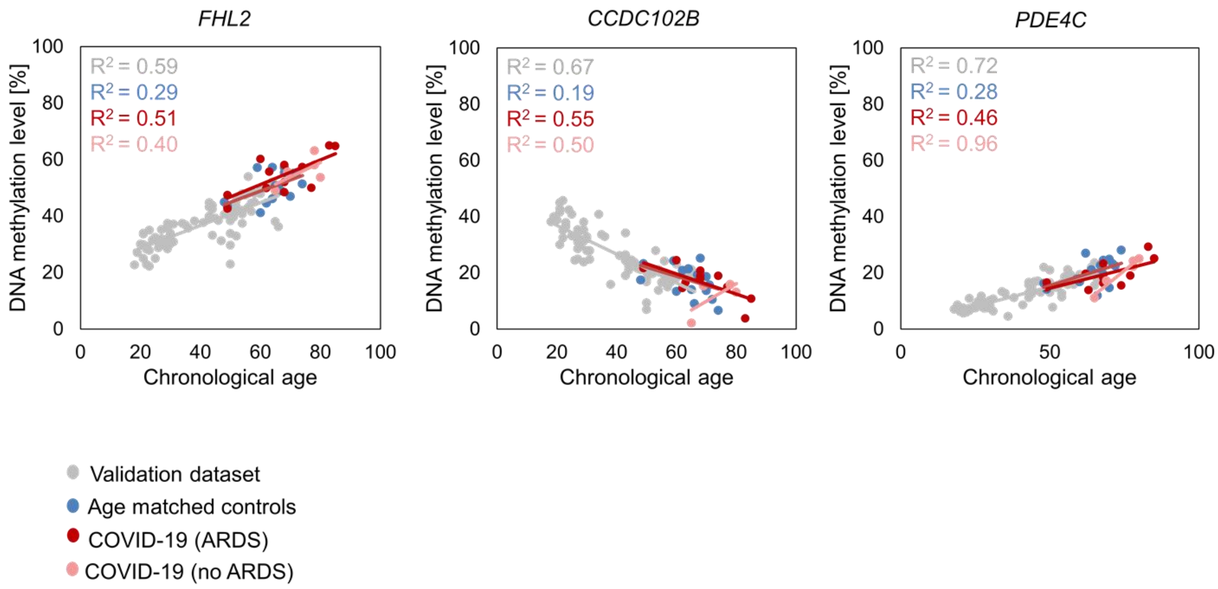


**Supplemental Figure S3. DNA methylation levels at the three age-associated CpGs.**

DNA methylation levels at the 3 CpG sites of our age predictor correlate with donor age. Samples of 17 COVID-19 patients with (n = 12, red) or without ARDS (n= 5, light red) do not show an offset in the DNA methylation levels in comparison to 17 age-matched healthy control samples (blue) or to the 78 healthy control samples, which were used for validation of the predictor (grey).

**Supplemental Table S1. Patient characteristics.**

| Patient ID | Gender | Age range* | Group | Outcome | EPIC | MiSeq | Flow-FISH |
|---|---|---|---|---|---|---|---|
| Patient 1 | M | 80 - 90 | no ARDS | dead | x | x | x |
| Patient 2 | M | 60 - 70 | ARDS | dead | x | x | |
| Patient 3 | F | 80 - 90 | ARDS | alive | x | x | x |
| Patient 4 | F | 70 - 80 | ARDS | alive | x | x | x |
| Patient 5 | M | 40 - 50 | ARDS | alive | x | x | x |
| Patient 6 | M | 40 - 50 | ARDS | dead | x | x | x |
| Patient 7 | M | 60 - 70 | ARDS | alive | x | x | x |
| Patient 8 | M | 60 - 70 | ARDS | dead | x | x | x |
| Patient 9 | F | 30 - 40 | no ARDS | alive | x | | x |
| Patient 10 | M | 60 - 70 | ARDS | alive | | x | x |
| Patient 11 | M | 60 - 70 | ARDS | alive | | | x |
| Patient 12 | F | 70 - 80 | ARDS | dead | | x | x |
| Patient 13 | M | 60 - 70 | no ARDS | alive | | x | x |
| Patient 14 | M | 80 - 90 | ARDS | alive | | x | x |
| Patient 15 | F | 60 - 70 | ARDS | dead | | x | x |
| Patient 16 | M | 70 - 80 | no ARDS | alive | | x | x |
| Patient 17 | F | 60 - 70 | ARDS | alive | | x | x |
| Patient 18 | F | 60 - 70 | no ARDS | dead | | x | x |
| Patient 19 | M | 70 - 80 | no ARDS | alive | | x | x |
| Patient 20 | M | 40 - 50 | no ARDS | alive | | | x |

* Precise age is not provided to prevent patient identifying information.

### Supplemental Methods

Blood samples of COVID-19 patients were taken at University Hospital of RWTH Aachen. All patient samples were taken after written and informed consent according to the guidelines and specific approval of the study by the local ethics committee (Ethic approval number EK 080/20 for the Covid-19 Aachen study named COVAS; Ethics committee of RWTH Aachen University, University Hospital Aachen, Pauwelsstrasse 30, 52074 Aachen, Germany) and collected into RWTH cBMB, the central biobank of the medical faculty of RWTH Aachen University (Ethic approval number EK 206/09). Blood samples of healthy donors were taken after written and informed consent according to the guidelines and approval of the study by the local ethics committee (EK 041/15; Ethics committee of RWTH Aachen University, University Hospital Aachen, Pauwelsstrasse 30, 52074 Aachen, Germany).

#### Analysis of DNA methylation microarray data

Genomic DNA of COVID-19 blood samples was isolated using the Maxwell 16LEV DNA Blood Kit (Promega, AS1290) in a Maxwell 16 instrument. 1200 ng DNA were bisulfite converted and analyzed with Illumina EPIC BeadChip microarrays at Life&Brain (Bonn, Germany). For comparison we exemplarily selected DNA methylation profiles of 185 peripheral blood samples (50 samples of GSE42861, 50 samples of GSE61496, 50 samples of GSE125105, and 35 samples of GSE123914) from Gene Expression Omnibus (https://www.ncbi.nlm.nih.gov/geo).

The idat files of the Illumina BeadChip datasets were quantile normalized using the R package minfi (Fortin et al., 2017). Principal component analysis was performed with the R package stats. Age predictions with the signatures of Horvath (2013) and Hannum et al. (2013) were performed with the R package wateRmelon (Pidsley et al., 2013). The more recent skin and blood clock of Horvath was estimated as described in Horvath et al. (2018). Age predictions of Han were performed with DNA methylation levels of 65 CpGs as previously described (Han et al., 2020). Predictions of leukocyte subsets were produced with the estimateCellCounts function of the R package minfi (Houseman et al., 2012; Jaffe and Irizarry, 2014).

#### Bisulfite amplicon sequencing

DNA methylation levels at three age-associated CpG sites (*FHL2, CCDC102B, PDE4C*) were analyzed by bisulfite amplicon sequencing as described in detail before (Franzen et al., 2017; Han et al., 2020). In brief, genomic DNA of 17 COVID-19 and 95 healthy control samples (17 age-matched controls and 78 controls of the validation set) was bisulfite converted using the EZ DNA methylation kit (Zymo Research). The three relevant genomic regions were amplified by PCR using the PyroMark PCR kit (Qiagen) and primers as described in Han et al. (2020) (Table S8 of that study). Illumina adapters were added by a second PCR and samples were sequenced on an Illumina MiSeq sequencer in 250 bp paired end mode using the V2 nano kit (Illumina). Mean read coverage was 3,370 reads for all amplicons and all samples. Alignment of reads to the hg19 genome build and calculation of methylation levels were performed with bismark (Krueger and Andrews, 2011).

#### Fluorescence in situ hybridization (flow-FISH)

Flow-FISH analysis of telomere length was carried out as previously described (Ferreira et al., 2020; Kirschner et al., 2020). Briefly, vital frozen mononuclear cells from peripheral blood were mixed with a FITC labeled telomere specific (CCCTAA)3-peptide nucleic acid FISH probe (Eurogentec) for DNA-hybridization followed by DNA counterstaining with LDS 751 (Sigma). Bovine thymocytes were used as internal controls. All measurements were carried out in triplicates. Telomere length is given in kilobases (Kb).
